# Impact of automated data flow and reminders on adherence and resource utilization for remotely monitoring physical activity in individuals with stroke or chronic obstructive pulmonary disease

**DOI:** 10.1101/2024.04.15.24305852

**Authors:** Margaret A. French, Aparna Balasubramanian, Nadia N. Hansel, Sharon K. Penttinen, Robert Wise, Preeti Raghavan, Stephen T Wegener, Ryan T. Roemmich, Pablo A. Celnik

## Abstract

As rehabilitation advances into the era of digital health, remote monitoring of physical activity via wearable devices has the potential to change how we provide care. However, uncertainties about patient adherence and the significant resource requirements needed create challenges to adoption of remote monitoring into clinical care. Here we aim to determine the impact of a novel digital application to overcome these barriers. The Rehabilitation Remote Monitoring Application (RRMA) automatically extracts data about physical activity collected via a Fitbit device, screens the data for adherence, and contacts the participant if adherence is low. We compare adherence and estimate the resources required (i.e., time and financial) to perform remote monitoring of physical activity with and without the RRMA in two patient groups. Seventy-three individuals with stroke or chronic obstructive pulmonary disease completed 28 days of monitoring physical activity with the RRMA, while 62 individuals completed 28 days with the data flow processes being completed manually. Adherence (i.e., the average percentage of the day that the device was worn) was similar between groups (p=0.85). However, the RRMA saved an estimated 123.8 minutes or $50.24 per participant month when compared to manual processes. These results demonstrate that automated technologies like the RRMA can maintain patient adherence to remote monitoring of physical activity while reducing the time and financial resources needed. Applications like the RRMA can facilitate the adoption of remote monitoring in rehabilitation by reducing barriers related to adherence and resource requirements.

## INTRODUCTION

Remote monitoring is an expanding area of digital health that allows healthcare professionals to obtain data about patients outside of the clinical environment, leading to reduced health disparities(1, 2), personalized(3) and preventative(4) medicine, value-based care(2, 5), and improved patient experiences(2) and outcome tracking(4). Remote monitoring is common in some medical disciplines (e.g., monitoring of blood glucose(6) or heart rhythm(7)) but not in rehabilitation, despite its potential to measure real-world function – the central target of rehabilitation.

Physical activity is one important aspect of function targeted during rehabilitation due to its association with hospital admission(8), risk of stroke(9), development of cardiovascular diseases(10, 11), and mortality(12–14). Physical activity is easily measured using wearable devices (e.g., Fitbits); however, clinical utility is limited by the time required to set up the device, download and analyze data, and synthesize, document, and discuss the results(2, 5, 15–17). These barriers are exacerbated by the lack of research demonstrating the value of remote monitoring approaches(3, 15, 17–19) and concerns about patient adherence(20).

We developed a Rehabilitation Remote Monitoring Application (RRMA) to address these barriers and advance remote monitoring approaches toward clinical implementation. Here we 1) compare patient adherence to physical activity remote monitoring with and without the RRMA and 2) estimate the resources saved when using the RRMA for physical activity remote monitoring. We hypothesize that the RRMA will maintain participant adherence while reducing the resources needed to collect data from wearable devices.

## METHODS

### Participants

One hundred thirty-five individuals (chronic obstructive pulmonary disease (COPD)=58; stroke=77) participated. They were in the MANUAL (n=62) or APP (n=73) group based on whether they enrolled before or after the launch of the RRMA (i.e., participants were not randomized; Figure 1). Inclusion criteria were smartphone ownership and in-home Wi-Fi; exclusion criteria included wheelchair use for mobility. Participants provided oral or written consent as approved by the Johns Hopkins University Institutional Review Board.

**Figure 1.**
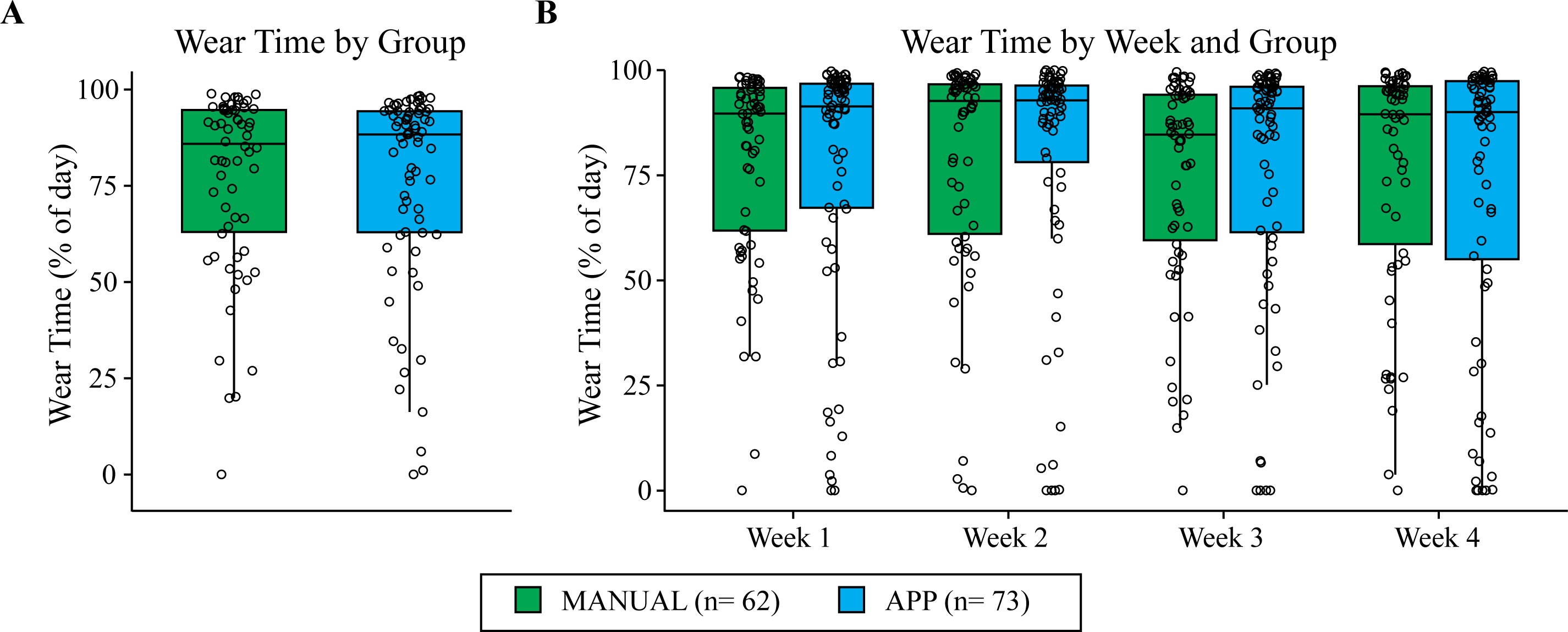
Consort diagram for the MANUAL and APP groups. Group assignment was based on time of enrollment.

### Procedures

We used Fitbit Inspire 2 (Fitbit Inc., San Francisco, California, USA) devices to measure minute-level step count and heart rate because they are widely accepted by patient populations(21–25) and data can be easily obtained through an Application Programming Interface (API). Participants were instructed to wear the devices at all times (except when showering or charging) on their non-dominant wrist (COPD) or their unimpaired wrist (stroke). Participants with stroke wore the device on their impaired wrist if hemiparesis limited their ability to place it on their unimpaired wrist. We asked participants to synchronize their Fitbit with their smartphone at least once/day.

### Data Management

The three components of data management included: 1) data extraction, 2) screening for participant adherence, and 3) providing reminders to synchronize devices. In the MANUAL group, we extracted Fitbit data using a custom-built Python script run every two weeks (Figure S1). We monitored participant adherence daily by logging into participants’ Fitbit accounts and documenting the last date of synchronization. If the participant had not synchronized their device in 5, 12, or 19 days, we attempted to contact them.

In the APP group, the RRMA (a Spring Boot application with an angular front end that runs on AKS Cluster) executed these three components automatically (Figure S1). The RRMA used study management information (e.g., date of enrollment, enrollment status) from Research Electronic Data Capture (REDCap)(26, 27) to ensure that it performed tasks for actively enrolled participants. The study team created and linked a unique, de-identified Fitbit account to the RRMA for each participant. This one-time linkage authorized the Fitbit API to share data with the RRMA. Once linked, the RRMA extracted minute-level data from the Fitbit API three times/day until the participant completed the study or withdrew. The RRMA then identified the most recent date that data was received. If data was not received in the past 5 or 12 days, the RRMA sent the participant a reminder to synchronize their device. If the device still was not synchronized after 19 days, the RRMA notified the study team to prompt human contact with the participant. The RRMA logged all reminders.

### Adherence

Adherence was defined by Wear Time, or the average percentage of the day during which a heart rate was detected. We used the accelerometry package(28) in R(29) to label each minute as worn or not worn and then calculated the percentage of each day that the device was worn.

We calculated Wear Time for each participant over 28-days and during each week individually. We used an independent t-test to compare Wear Time during the entire time period between groups (MANUAL vs. APP) and a 4 (week) x 2 (group) mixed effects ANOVA to compare across weeks and between groups. We used α≤0.05 for all analyses.

### Resource Utilization

Total Cost related to time - the sum of resources required to complete the three data management steps outlined above - quantified resource utilization. We conservatively estimate that manual data extraction and adherence screening each takes 2 minutes/day/participant. The RRMA automated data extraction and adherence screening, but the time needed to manually link the RRMA and Fitbit accounts was included as time required for data extraction.

To estimate the time needed to provide synchronizing reminders, we used the RRMA’s reminder log to calculate the average number of reminders/participant month. We used the average number of all reminders (i.e., 5-, 12-, and 19-day) to determine the time needed to provide reminders manually at each time point. The RRMA automated the 5- and 12-day reminders; thus, we used the number of 19-day reminders/participant month to determine the time needed to provide reminders with the RRMA. This resulted in the use of 0.98 and 0.1 reminders/participant month in the Total Cost calculations without and with the application, respectively. We conservatively estimated that each manual reminder takes 10 minutes.

We used the Total Cost of time to calculate a secondary metric of Total Cost related to money. To convert the resources related to time to money, we assumed a $50,000 annual salary for a study coordinator working 40 hours/week to complete these tasks. We present Total Cost related to time and money per participant month and the resources saved by using the RRMA, calculated as the difference between Total Cost with and without the RRMA.

## RESULTS

There were no significant differences in demographic characteristics between the MANUAL and APP groups with the exception of race (Table 1). Demographic information based on diagnosis is in Table S1. Diagnosis-specific clinical characteristics are shown in Table 2.

**Table 1.** Demographic information.

| **Characteristic** | **Excluded<sup>a</sup>**<br>N = 41 | **Included<sup>a</sup>**<br>N = 135 | **p-value<sup>b</sup>** | **Manual<sup>a</sup>**<br>N = 62 | **App<sup>a</sup>**<br>N = 73 | **p-value<sup>b</sup>** |
| --- | --- | --- | --- | --- | --- | --- |
| **Age** | 63 (13) | 64 (11) | 0.7 | 66 (9) | 63 (12) | 0.12 |
| **Race** |  |  | 0.8 |  |  | <0.001 |
| White | 23 (56.1%) | 80 (59%) |  | 48 (77.4%) | 32 (43.8%) |  |
| Black | 16 (39%) | 45 (33%) |  | 13 (21%) | 32 (43.8%) |  |
| Other | 2 (4.9%) | 10 (7.4%) |  | 1 (1.6%) | 9 (12.3%) |  |
| **Sex: Female** | 28 (68.3%) | 64 (47.4%) | 0.019 | 31 (50%) | 33 (45.2%) | 0.6 |
| **Ethnicity** |  |  | 0.3 |  |  | 0.2 |
| Hispanic | 2 (4.9%) | 2 (1.5%) |  | 0 (0%) | 2 (2.7%) |  |
| Not Hispanic | 39 (95.1%) | 133 (98.5%) |  | 62 (100%) | 71 (97.3%) |  |

<sup>a</sup> Mean (SD); n (%)
<sup>b</sup> Wilcoxon rank sum test; Fisher's exact test; Pearson's Chi-squared test

**Table 2.** Diagnosis specific clinical information.

| Characteristic | Excluded<sup>a</sup> | Included<sup>a</sup> | p-value<sup>b</sup> | Manual<sup>a</sup> | App<sup>a</sup> | p-value<sup>b</sup> |
| --- | --- | --- | --- | --- | --- | --- |
| **Stroke** |  |  |  |  |  |  |
| **N** | 20 | 77 |  | 9 | 68 |  |
| **Time since stroke (mo)** | 8 (16) | 30 (51) | 0.02 | 42 (74) | 28 (48) | 0.5 |
| **Affected side: Left** | 6 (30%) | 45 (58.4%) | 0.02 | 5 (56%) | 40 (59%) | >0.9 |
| **Assistive Device** |  |  | >0.9 |  |  | >0.9 |
| None | 12 (60%) | 44 (57%) |  | 5 (56%) | 39 (57%) |  |
| Cane | 6 (30%) | 24 (31%) |  | 3 (33%) | 21 (31%) |  |
| Walker | 2 (10%) | 9 (12%) |  | 1 (11%) | 8 (12%) |  |
| **COPD** |  |  |  |  |  |  |
| **N** | 21 | 58 |  | 53 | 5 |  |
| **History of Smoking: Yes** | 20 (95%) | 55 (95%) | >0.99 | 50 (94%) | 5 (100%) | >0.99 |
| **FEV1 % predicted** | 61 (21) | 52 (20) | 0.2 | 52 (20) | 55 (48) | 0.9 |
| **FEV1/FVC ratio** | 63 (15) | 57 (14) | 0.12 | 58 (13) | 48 (12) | 0.12 |
| **Hospitalization during year prior to enrollment: Yes** | 17 (81%) | 46 (79%) | >0.99 | 41 (77%) | 5 (100%) | 0.6 |

<sup>a</sup>Mean (SD); n (%)
<sup>b</sup>Wilcoxon rank sum test; Fisher's exact test; Pearson's Chi-squared test

### Adherence

Wear Time during the entire 28-day period was not significantly different between the MANUAL and APP groups (MANUAL=75.8±26.0%, APP=76.7±23.5%, p=0.85; Figure 2A).

**Figure 2.**
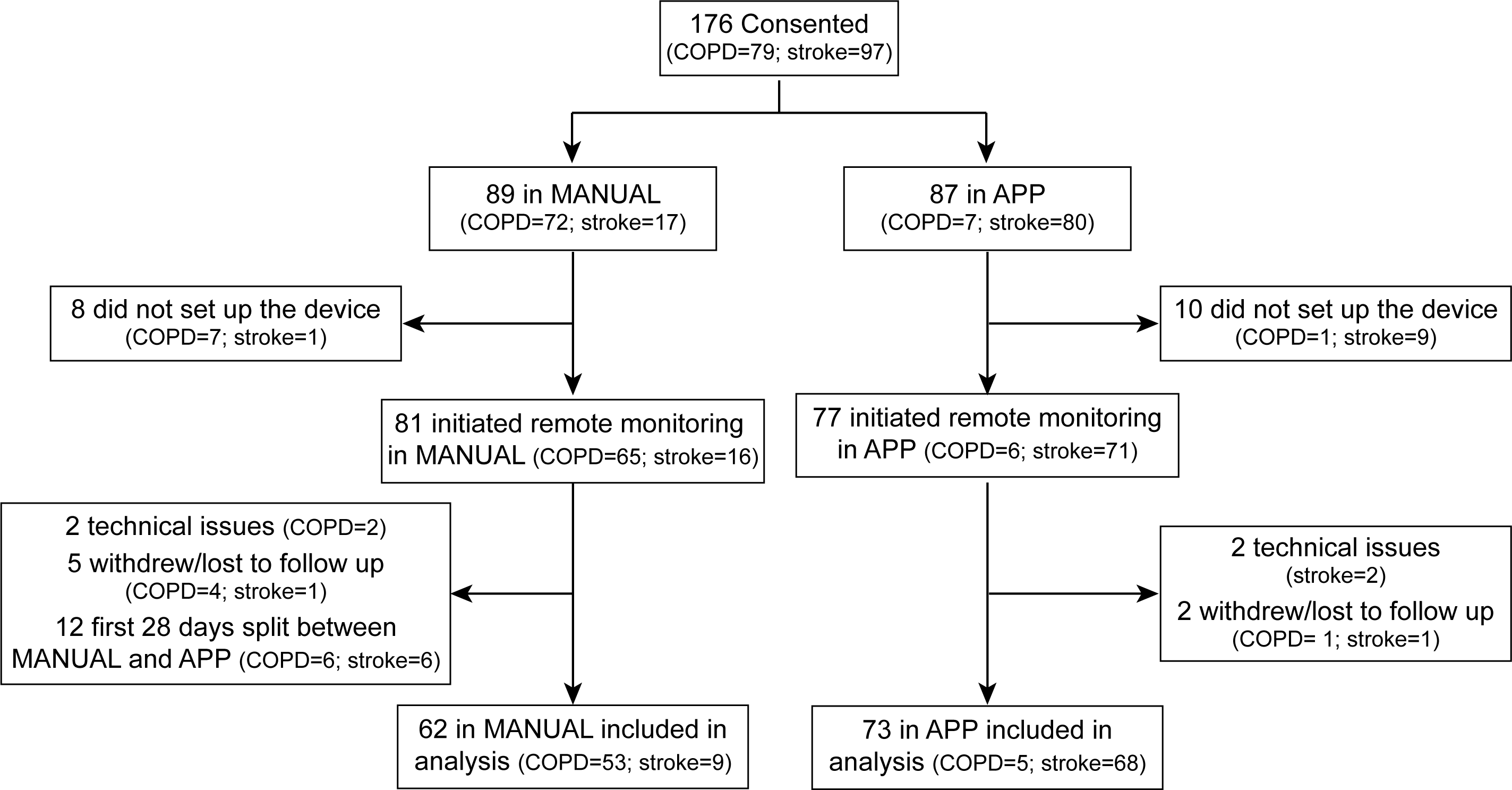
Adherence as measured by Wear Time in the MANUAL and APP groups during the full 28-day period (A) and during each week of the 28-day period (B).

As a sensitivity analysis, we performed the same analysis with the seven individuals who withdrew during their first 28 days, showing no significant difference between groups (p=0.78). There was no significant main effect of week (F_(2.3,303.8)_=2.1, p=0.12) or group (F_(1,132)_=0.04, p=0.8) on Wear Time and no significant week x group interaction (F_(2.3,303.8)_=0.81, p=0.46; Figure 2B), indicating that overall adherence and adherence over time were not impacted by the RRMA. The impact of diagnosis was beyond the scope of this study; however, we show these data in Figure S2.

### Resource Utilization

Total Cost of time without and with the RRMA was 125.8 and 2 minutes/participant month, respectively (Table 3). We estimate that this process costs $50.32/participant month without the RRMA and $0.08/participant month with the RRMA, resulting in a savings of $50.32/participant month with the RRMA (Table 3). This would have resulted in $81,518.4 savings if our population would have completed a yearlong study.

**Table 3.** Resource utilization without and with the RRMA.

|  | Time (minutes per participant month)<sup>a</sup> |  |  | Money (US dollars per participant month)<sup>a</sup> |  |  |
| --- | --- | --- | --- | --- | --- | --- |
|  | Without RRMA | With RRMA | Resources saved | Without RRMA | With RRMA | Resources saved |
| **Data Extraction** | 58 | 1<sup>b</sup> | 57 | 23.20 | 0.04<sup>b</sup> | 23.16 |
| **Adherence Screens** | 58 | 0 | 58 | 23.20 | 0 | 23.40 |
| **Synchronization Reminders<sup>c</sup>** | 9.8 | 1 | 8.8 | 3.92 | 0.04 | 3.88 |
| **Total Cost** | 125.8 | 2 | 123.8 | 50.32 | 0.08 | 50.24 |

Abbreviations: RRMA- Rehabilitation remote monitoring application.
<sup>a</sup> 28 days was considered one month
<sup>b</sup> Reflects the need to manually link the participant's Fitbit account to the RRMA. This time is only needed during the first month of enrollment.
<sup>c</sup> Estimates of time and money needed to provide reminders without the RRMA are based on the average number of all reminders (i.e., 5-, 12-, and 19-day) needed per participant month. With the RRMA, the 5- and 12-day reminders are provided via email or text; thus, only the 19-day reminders are considered in the condition with the RRMA.

## DISCUSSION

Wearable devices can improve how we measure physical activity clinically, but barriers such as patient adherence and time and resource requirements impede our use of these approaches. Here we demonstrate that technology (in the form of the RRMA) can address these barriers by maintaining patient adherence while drastically reducing the resources needed to obtain quality data from remote monitoring.

Adherence to remote monitoring is critical for scalability into clinical care. We found high adherence to monitoring physical activity, which is consistent with past work in several patient groups (30–34). Importantly, our findings demonstrate that adherence is similar when reminders are placed manually or automatically. This shows that automated technologies like the RRMA can manage adherence, drastically improving the scalability of remote monitoring.

The time and money needed to extract data from Fitbit devices, screen this data for adherence, and remind participants to synchronize their devices was markedly reduced by the RRMA. The resources needed to manually perform these tasks are likely not feasible in a clinical setting (15). Our work suggests that adherence is maintained when utilizing a less resource intensive and automated process, providing an avenue for improved feasibility of remote monitoring in clinical care.

We acknowledge some limitations. The duration of clinical care in rehabilitation is often longer than 28 days as studied here, and we only used one reminder schedule. There is likely a tradeoff between the frequency of reminders, resource utilization, and adherence that should be explored. Lastly, studies with prospective randomization would provide valuable insight into how adherence is impacted by automated reminders.

## DECLARATIONS

### Conflicting interests

The Authors declare that there is no conflict of interest.

## Funding

This work was supported by the Sheikh Khalifa Stroke Institute; the Johns Hopkins’ inHealth Precision Medicine Initiative; and the National Institutes of Health [grant number 1F32HD108835-01].

## Ethical approval

The Institutional Review Board of Johns Hopkins University approved this study (IRB00247292 and IRB00236214).

## Guarantor

RR

## Contributorship

MF, RR, NH, SW, and PC conceived the study and participated in protocol development. MF, PR, BW, SP, and NH were involved in gaining approval from the Institutional Review Board. MF, AB, BW, and NH were involved in participant recruitment. MF and RR performed data analysis and data interpretation. All authors contributed to the interpretation of the data. MF wrote the first draft of the manuscript. All authors reviewed and edited the manuscript and approved the final version of the manuscript.

## Data Availability

All data produced in the present study are available upon reasonable request to the authors.

## Acknowledgements

The authors acknowledge the Johns Hopkins Technology Innovation Center for the development of the application described in this work.

## Notes

### Competing Interest Statement

The authors have declared no competing interest.

### Author Declarations

The data in this work was collected under an approved the Johns Hopkins University Institutional Review Board, protocol numbers IRB00236214 and IRB00247292.

