## Supplementary material for "Impact of automated data flow and reminders on adherence and resource utilization for remotely monitoring physical activity in individuals with stroke or chronic obstructive pulmonary disease": Table S1; Figure S1; Figure S2

Table S1. Demographic information by diagnosis

|  | **Inclusion Status** | | | | | | | | **Group Membership** | | | | | | | | |
| --- | --- | --- | --- | --- | --- | --- | --- | --- | --- | --- | --- | --- | --- | --- | --- | --- | --- |
|  | **COPD** | | | | **Stroke** | | | | **COPD** | | | | | **Stroke** | | | |
| **Characteristic** | **Excluded^1^**  N = 21 | **Included**^1^  N = 58 | **p-value**^2^ | **Excluded^1^**  N = 20 | | **Included**^1^  N = 77 | **p-value**^2^ | **MANUAL**^1^  N = 53 | | **APP**^1^  N = 5 | **p-value**^2^ | | **MANUAL**^1^  N = 9 | | **APP**^1^  N = 68 | | **p-value**^2^ |
| **Age** | 69 (12) | 68 (8) | 0.3 | 57 (11) | | 62 (12) | 0.12 | 67 (8) | | 69 (10) | 0.6 | | 59 (13) | | 62 (12) | | 0.4 |
| **Race** |  |  | 0.3 |  | |  | 0.3 |  | |  | 0.1 | |  | |  | | 0.1 |
| Black | 4 (19%) | 15 (26%) |  | 12 (60%) | | 30 (39%) |  | 12 (23%) | | 3 (60%) |  | | 1 (11%) | | 29 (43%) | |  |
| Other | 1 (4.8%) | 0 (0%) |  | 1 (5.0%) | | 10 (13%) |  | 0 (0%) | | 0 (0%) |  | | 1 (11%) | | 9 (13%) | |  |
| White | 16 (76%) | 43 (74%) |  | 7 (35%) | | 37 (48%) |  | 41 (77%) | | 2 (40%) |  | | 7 (78%) | | 30 (44%) | |  |
| **Sex: Female** | 17 (81%) | 32 (55%) | 0.037 | 11 (55%) | | 32 (42%) | 0.3 | 29 (55%) | | 3 (60%) | >0.9 | | 2 (22%) | | 30 (44%) | | 0.3 |
| **Ethnicity** |  |  | 0.3 |  | |  | 0.4 |  | |  | >0.9 | |  | |  | | >0.9 |
| Hispanic | 1 (4.8%) | 0 (0%) |  | 1 (5.0%) | | 2 (2.6%) |  | 0 (0%) | | 0 (0%) |  | | 0 (0%) | | 2 (2.9%) | |  |
| Not Hispanic | 20 (95%) | 58 (100%) |  | 19 (95%) | | 75 (97.4%) |  | 53 (100%) | | 5 (100%) |  | | 9 (100%) | | 66 (97%) | |  |
| ^1^Mean (SD); n (%) | | | | | | | | | | | | | | | |  |  |
| ^2^Wilcoxon rank sum test; Fisher's exact test; Pearson's Chi-squared test | | | | | | | | | | | |  | | | |  |  |

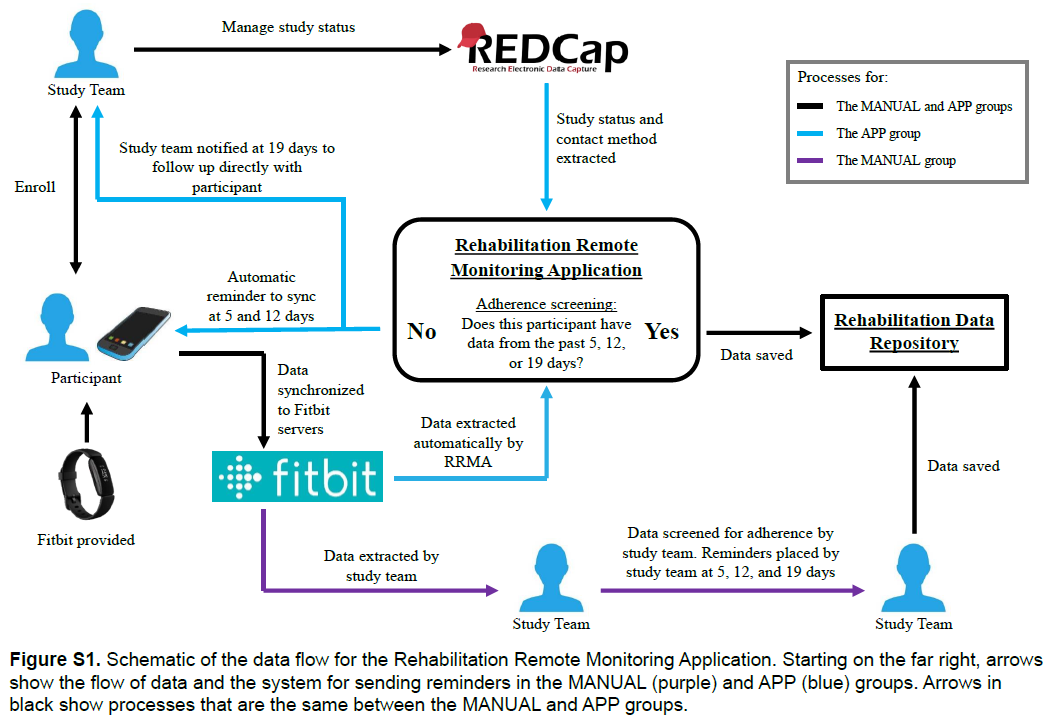

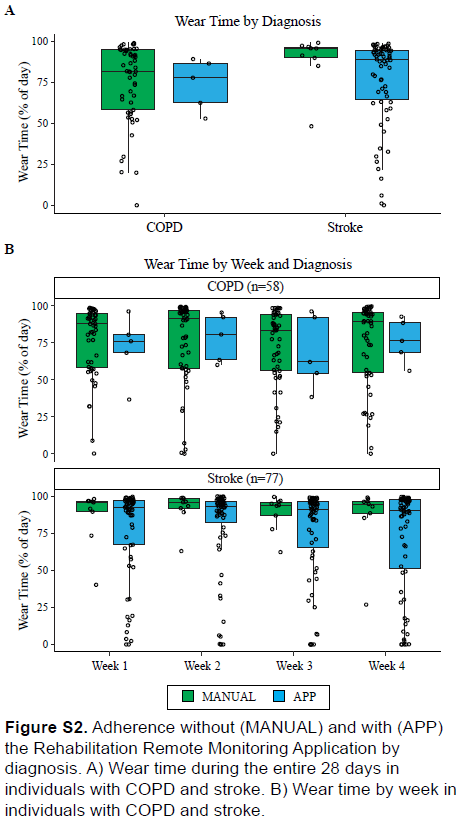
